# MRI versus mammography plus ultrasound in women at intermediate breast cancer risk: study design and protocol of the MRIB multicentre randomized controlled trial

**DOI:** 10.1101/2021.03.10.21253341

**Authors:** Luigina Ada Bonelli, Massimo Calabrese, Paolo Belli, Stefano Corcione, Claudio Losio, Stefania Montemezzi, Federica Pediconi, Antonella Petrillo, Chiara Zuiani, Lucia Camera, Luca Alessandro Carbonaro, Andrea Cozzi, Daniele De Falco Alfano, Licia Gristina, Marta Panzeri, Ilaria Poirè, Simone Schiaffino, Simona Tosto, Giovanna Trecate, Rubina Manuela Trimboli, Francesca Valdora, Sara Viganò, Francesco Sardanelli

## Abstract

**Background:** In women at high/intermediate lifetime risk of breast cancer (BC-LTR), contrast-enhanced magnetic resonance imaging (MRI) added to mammography ±ultrasound (Mx±US) increases sensitivity but decreases specificity. An alternative strategy, MRI alone, potentially more cost-effective, has never been explored. This study aims to assess the characteristics of women who participated in a randomized trial offering MRI alone.

**Methods:** In this feasibility multicentre randomized controlled trial we compared MRI alone versus Mx+US in women at intermediate BC-risk (allocation ratio 1:1). Eligible women were aged 40 –59, with a 15–30% LTR and/or extremely dense breasts. Two screening rounds per woman were planned in ten centres experienced in MRI screening. Primary endpoint: rate of cancers detected in the two arms after 5 years of follow-up. Secondary endpoints: distribution of the risk profiles among the women enrolled in the trial; distribution of pathological stages and histology of cancers detected; interval cancer rate in the two arms.

**Results:** From 07/2013 to 11/2015, 1,254 women (mean age 47 years) were enrolled: 624 were assigned to Mx+US, 630 to MRI. Most of them were aged below 50 (72%) and premenopausal (45%), 52% used oral contraceptives. Among postmenopausal women, 15% had used hormone replacement therapy. Breast and/or ovarian cancer in mother and/or sisters were reported by 37% of enrolled women, extreme breast density was recorded for 79%, 41% had a 15–30% BC-LTR.

**Conclusions:** The distribution of BC-risk profile major determinants (breast density and family of breast and ovarian cancer) of enrolled women varied across centres.

**Trial registration:** NCT02210546

**Key points:**

- Mammography plus ultrasound are commonly used to screen women with dense breasts
- Supplemental ultrasound increases BC detection rate but also false-positives and potential overdiagnosis
- Whether increased BC detection by US translates into reduced mortality is unknown
- MRI alone could be more risk- and cost-effective than mammography plus ultrasound

## Background

Mammography (Mx) represents the primary screening tool for breast cancer (BC), but its preventive impact is not fully satisfactory. Considering the screening age range, Mx yields an estimated BC mortality reduction of about 30% in the target population, with a beneficial effect persisting for at least ten years [1, 2]. However, even for women who regularly adhere to a screening program, risk reduction brought about by Mx screening remains approximately 40% [3]. Such limited efficacy has been attributed not only to the intrinsic limitations of Mx but also to the highly variable biological characteristics of BC [4], as well as to women’s individual characteristics such as age and breast density (BD). A high BD is not only an independent BC risk factor for both premenopausal and postmenopausal women [5, 6], but also reduces Mx sensitivity (masking effect), resulting in an increased interval cancer rate [7–9]. While tomosynthesis, contrast-enhanced magnetic resonance imaging (MRI) and contrast-enhanced mammography may all display an increased sensitivity compared to Mx, especially for small BCs [10–12], a more sensitive test could detect more small tumours only because they are growing slowly, potentially leading to an increase in overdiagnosis [13].

In high-risk women, the addition of Mx to MRI did not substantially increase sensitivity [14–18]. Moreover, supplementary screening breast ultrasound (US) or MRI in addition to Mx in these women resulted in a higher cancer detection rate [16, 19], but also increased the false-positive rate [16, 20]. In this context, any attempt to improve screening efficacy by increasing the sensitivity of the process/test(s) needs to be carefully evaluated, since it might produce more harms than benefits. Furthermore, no study has demonstrated that the addition of MRI to traditional imaging in high-risk women reduces BC-related mortality. In the absence of clear evidence, there is no consistent recommendation from international guidelines on a threshold of BC lifetime risk (BC-LTR) that warrants the recommendation of periodic MRI surveillance [21, 22].

In two cohort studies outside the high-risk setting, focused on healthy women who underwent MRI and Mx with or without US, MRI had better sensitivity than Mx with or without US, particularly in women with dense breasts [11, 23]. No interval cancers were observed [11], but the net benefit and additional costs of MRI were not estimated. In the DENSE trial, the supplemental MRI screening in women with in extremely dense breasts and negative Mx resulted in a significantly lower interval BC rate than Mx alone [24].

However, an add-on strategy (i.e., adding tests to Mx) to increase the sensitivity of the screening process may decrease specificity and potentially increase overdiagnosis. An alternative strategy, i.e., replacing Mx plus US with MRI, could be more risk-and cost-effective. This hypothesis has never been explored in a classical head-to-head trial. However, a typical efficacy trial requires the recruitment of tens of thousands of subjects and would require evidence, up to now unavailable, on the acceptability and actual performance of MRI as a stand-alone screening test among women at intermediate risk of BC are not available. Thus, a feasibility multicentre randomized controlled trial (RCT) aimed at comparing the performance of contrast-enhanced MRI as a stand-alone screening tool versus Mx plus US in women at intermediate BC risk was started in 2013 in Italy. Here we focus on a secondary endpoint of the study, i.e., the analysis of the distribution of patients’ characteristics and BC-risk profiles in the enrolled cohort.

## Methods

The study was in accordance with the ethical standards of the institutional research committees (Ethics Committee of Regione Liguria for the coordinating centre and each participating centre competent Ethics Committee). All women enrolled in the study received an information sheet and signed a written informed consent.

### Study design and population

Women were randomly assigned to receive annual 2D Mx plus US (standard of care arm) or MRI (experimental arm) with a 1:1 allocation ratio. Two screening rounds per woman were planned.

Women were deemed eligible for enrollment if aged 40 –59 years and if they had a 15–30% BC-LTR and/or extreme BD on the most recent Mx. BC-LTR was calculated using the IBIS Breast Cancer Risk Evaluation Tool version 6.0.0 (http://www.ems-trials.org/riskevaluator/).

Exclusion criteria were: signs or symptoms of BC, previous BC (invasive or ductal in situ), cancer at any other site, presence of life-threatening diseases, known *BRCA* or *TP53* pathogenic germ-line mutation carriers, pregnancy. We also excluded: women with general contraindications to MRI or to intravenous administration of gadolinium-based contrast agent; women who underwent hormonal enhancement of ovarian function for medically assisted reproduction in the previous three years; women planning a pregnancy; women undergoing postmenopausal hormone replacement therapy (HRT) who refused to suspend the treatment four weeks before MRI performance.

### Enrolment

Women aged 40–59 years who had an Mx scheduled during the study period were interviewed to check their eligibility. They were informed about the study aims and about the associated potential risks and benefits. Those who accepted to participate signed an informed consent and were randomized. Randomization was centralized via web-based connection to the coordinating centre (http://ctrials.hsanmartino.it/ist/rde/). After eligibility had been checked, the assignment of each woman was disclosed to the centre. Randomization was stratified according to centre and women age at enrolment (< 50 or ≥ 50 years).

### Participating centres and imaging readers

According to the EUSOMA recommendations [25], the following facilities had to be available at each participating centre: a) an electronic image storage system for Mx, US, and MRI; b) full-field digital Mx systems; c) breast US scanner; d) MR units with magnets with field intensity ≥ 1.0 T and gradients ≥ 20 mT/m (details are reported Table 1). Radiologists involved in the study had to provide documented experience in breast imaging, i.e., the performance and/or interpretation of at least 500 breast MRI, 10,000 Mx, and 5,000 US examinations, also documenting adequate skill in interventional procedures (under stereotactic or US guidance), and at least 150 breast MRI examinations performed in the previous year. In addition, participating centres had to guarantee: the performance of needle biopsy (core or vacuum-assisted) under stereotactic, US, and MRI guidance; second-look US and re-evaluation of Mx to identify MRI-detected lesions; availability of preoperative localization under stereotactic, US, and MRI guidance.

**Table 1.** General requirements for radiology departments to participate in the MRIB study

|  |  |
| --- | --- |
| **1** | Availability of electronic image storage system for Mx, US, and MRI |
| **2** | Full-field digital Mx systems with high-resolution electronic display systems available to both the technologist at the time of the examination and the interpreting physician; relevant information on the digital images and the patient examined available in the display settings of the dedicated workstation |
| **3** | Breast US scanner equipped with a multi-frequency linear array transducer operating at a centre frequency higher than 10 MHz |
| **4** | MR units with magnets with intensity field $\geq 1.0$ T and gradients $\geq 20$ mT/m, equipped with bilateral dedicated coils, preferably multichannel and an automated power injector system with double syringe for both contrast agent and normal saline solution. MRI protocol must include a high-contrast bilateral morphologic sequence and a bilateral dynamic two-dimensional or three-dimensional study with spatial in-plane resolution $\leq 1.5\text{mm}^2$ (preferably $\leq 1\text{ mm}^2$), temporal resolution $\leq 120$ s |

Ten qualified centres with long experience in breast MRI screening for high-risk women (as in the HIBCRIT study [15]) joined the study and a team of investigators was established at each centre.

### Imaging interpretation

Examinations were interpreted according to the BI-RADS classification system for Mx, US and MRI [26]: category 0 (non-diagnostic), category 1 (negative), category 2 (benign), category 3 (probably benign, i.e., cancer probability < 2%), category 4 (suspicious abnormality —biopsy should be considered) and category 5 (highly suggestive of malignancy —appropriate action should be taken).

Mammographic BD was evaluated by visual analysis and was categorized according to the following American College of Radiology (ACR) categories: 1) almost entirely fat (*a*), 2) scattered fibroglandular densities (*b*); 3) heterogeneously dense, which may obscure small masses (*c*); 4) extremely dense, which lowers Mx sensitivity (*d*) [26]. When the breasts were not of apparently equal BD, the denser one was used to categorize BD.

### Diagnostic workup

In both study arms, women with examinations classified as BI-RADS category 0 repeated imaging tests; those classified as BI-RADS categories 1 or 2 were returned to the assigned arm or to usual screening if the two study rounds had been completed. Examinations classified as BI-RADS category 4 or 5 were immediately invited to undergo diagnostic and interventional procedures as appropriate.

Women who had Mx and/or US classified as BI-RADS category 3 were invited to repeat Mx and/or US within 6 months (early recall) according to the characteristics of the detected lesion(s). If the early recall exam(s) was classified as BI-RADS category 1 or 2, the women returned to the assigned group or to screening; if it was classified as BI-RADS category 3 to 5, the women were invited to undergo diagnostic and interventional procedures.

Women who had MRI classified as BI-RADS category 3 were referred to second-look US and/or re-evaluation of Mx according to the characteristics of the observed lesion(s). Those who had the second-look/re-evaluation test(s) classified as BI-RADS category 1 or 2 were returned to the assigned study arm or to screening. Women with test(s) classified as BI-RADS category 3 were invited to repeat Mx and/or US after 3 months; short term follow up with MRI was not considered. If the result of the 3-month examination(s) remained BIRADS category 3, or worsened to category BI-RADS 4, or 5, the women were invited to undergo interventional procedures.

Diagnostic and interventional procedures performed in the work-up of detected abnormalities (either after screening examination or after short term follow up) included fine needle sampling, core needle biopsy with at least 14-G bore devices with or without coaxial systems, and vacuum-assisted biopsy with at least 11-G bore devices. The diagnostic workup in the two study arms is reported in Fig. 1.

**Fig. 1a.**
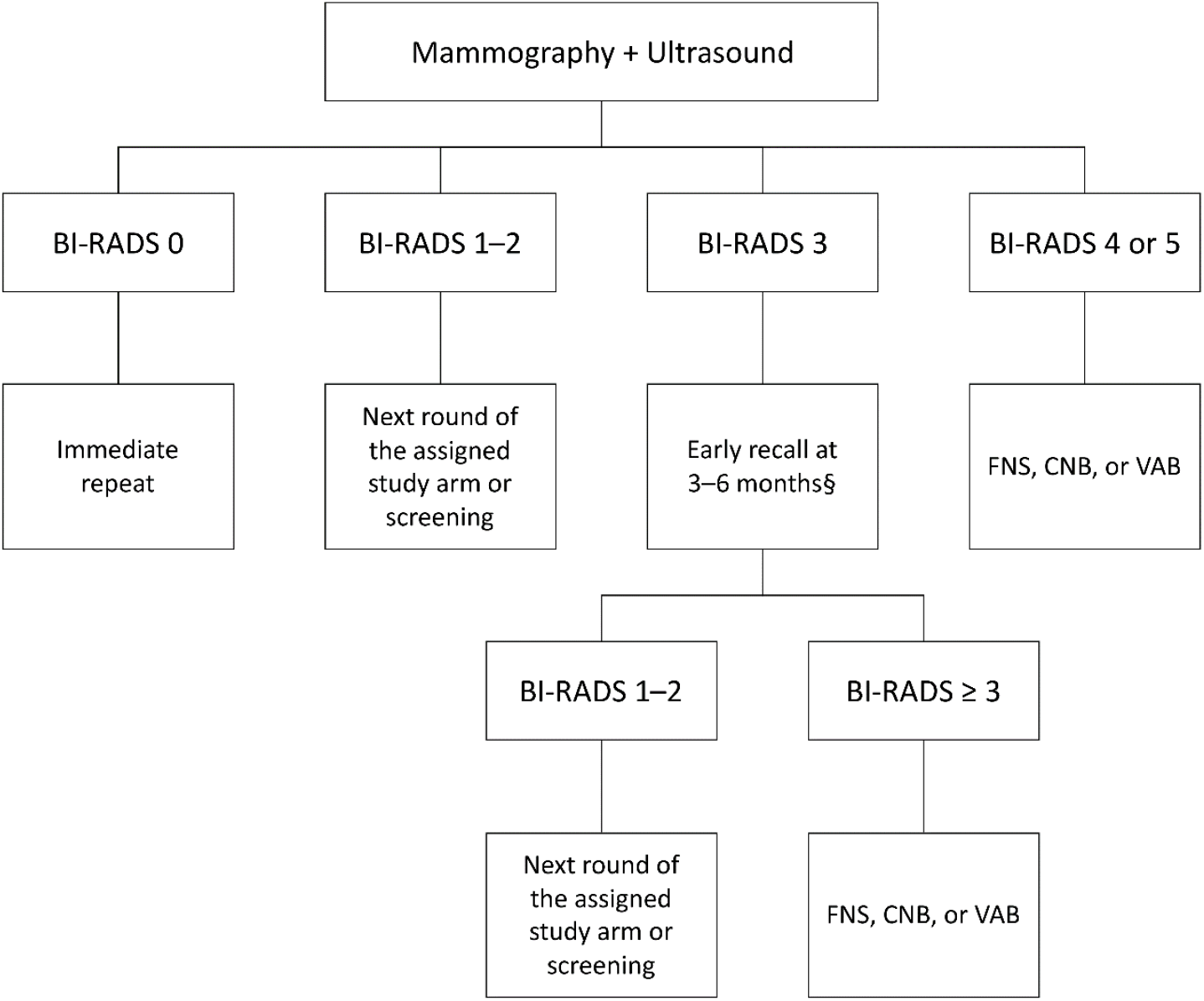
**a)** Diagnostic work up for women assigned to the *standard of care arm* with suspicious mammography and/or ultrasound

**Fig. 1b.**
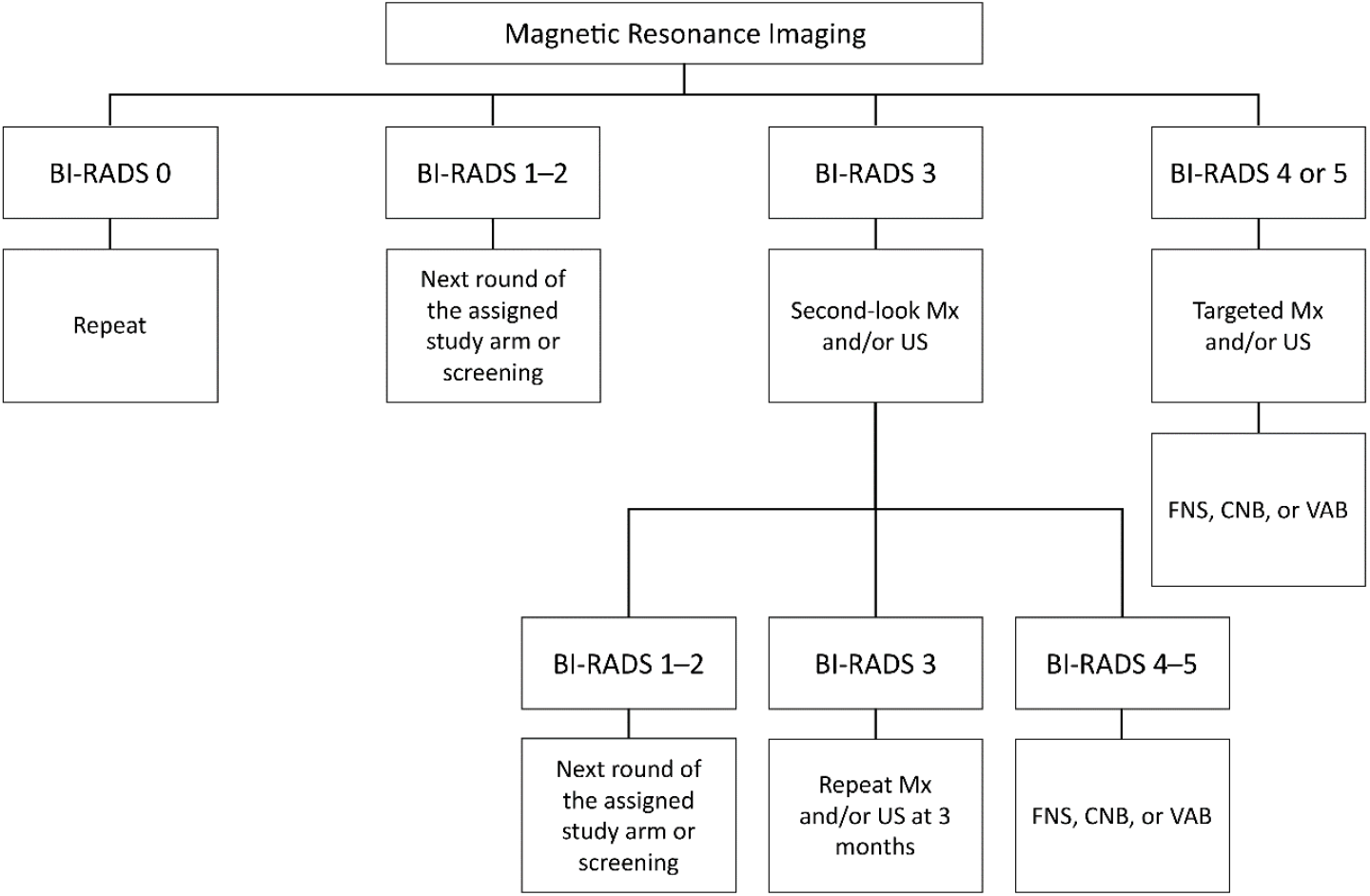
**b)** Diagnostic work up for women assigned to *experimental arm* with suspicious breast magnetic resonance imaging

### Data collection

The following data were collected upon enrolment of each woman: reproductive history, use of birth control pill, HRT, height, weight, alcohol consumption, tobacco smoking history, first-degree family history of BC and of ovarian cancer (OC), ACR density class at the most recent Mx, and BC-LTR.

The coordinating centre was responsible for data storage, monitoring, and quality controls of the study. as well as of the assessment of main study endpoints. The Clinical Trials Centre of the coordinating centre developed the e-case report forms to record: imaging data examinations, pathology data for lesions biopsied and/or removed, details of surgical procedures, stage of detected BC, eventual adverse events. A password-protected database was designed and managed, each researcher receiving a personal login/password. The Clinical Trials Centre was also in charge of monitoring data collection and auditing the filled-in case report forms. Participating centres provided de-identified data according to current regulations. At the Clinical Trials Centre, the enrolled women were identified with a unique study number assigned at randomization.

### Study endpoints

The primary endpoint of this study was the rate of invasive and ductal in situ BC detected in the two study arms. All BCs diagnosed as a consequence of an abnormality identified at the screening tests were considered screen-detected.

Secondary endpoints include: a) the distribution of clinical and pathological stages of invasive BCs; b) the histological characteristics of BCs; c) the interval cancer rate, both between the two examinations and within one year from the second examination: any BC (invasive or ductal in situ) diagnosed after a negative examination but before the following examination, scheduled approximately 1 year later, will be considered as an interval cancer; d) the adherence to the assigned arm and any reason for consent withdrawal from the assigned program; e) the distribution of the BC risk profile; f) the number of BCs (invasive and in situ) detected in excess in the experimental arm compared to the conventional arm (overdiagnosis) after 4 years of follow up. BC risk profile was built combining the LTR score (< 15% or ≥ 15%) and the BD at the most recent mammography before enrolment (ACR class *a* to *c*, or ACR class *d*), so that four risk categories (risk profile) were created: 1) LTR ≥ 15% and BD = *a* to *c*; 2) LTR < 15% and BD = *d*; 3) LTR ≥ 15% and BD = *d*; 4) LTR ≥ 15% and unknown BD.

### Sample size estimation

The study was designed as a feasibility study, preliminarily to the conduction of a large-size efficacy trial. The size of an efficacy trial with BC mortality (or incidence of metastatic BC) as the primary endpoint should allow to observe at least 380 events (BC deaths, or incidence of metastatic BC) in order to detect – with an 80% power – a 25% relative reduction in BC mortality, which is considered the minimal effect of MRI screening that is worth detecting. Assuming a 60% survival at 10 years, and an average cumulative 10-year BC risk of 5% (the lowest risk in this cohort should be about 3%), we can estimate that at least 20,000 women followed for 10 years (with a further follow-up of BCs until 380 events have been observed) would be necessary for such an efficacy trial. However, accrual for large-size prevention trials is very difficult, as they target asymptomatic healthy individuals facing a variable but generally low risk. Thus, this feasibility trial aimed to estimate the sensitivity and specificity of MRI screening alone, but also to provide information on the distribution of the risk profiles among enrolled women, as well as to estimate the sample size needed by an efficacy trial. Therefore, a planned enrolment of 2,000 women (10% of the size of the efficacy trial) was proposed. Furthermore, organizational problems and quality control issues could be adequately addressed in a study of this size.

It can be expected that in this feasibility trial, over a 5-year screening period, 40–60 cases of invasive BC will be observed: one third in the control arm; two thirds in the MRI arm. These figures approach those of previous uncontrolled MRI studies (e.g., the HIBCRIT study [15]), enabling to confirm the two-fold increase in sensitivity associated with MRI. These figures were also considered sufficient to provide preliminary information on the MRI-associated lead time, and on the stage distribution of incident BCs. Conversely, the number of advanced (metastatic or locally advanced) BCs and the number of BC-related deaths should be too small to allow any meaningful interpretation. Due to lead time, no effects of MRI on efficacy endpoints are expected to be noted during the first years. Any estimation of the proportion of interval cancers in each arm proved to be difficult, being this proportion dependent on the age distribution of enrolled women.

Two and half years since the start of the study, when 1,254 women had been randomized, enrollment stopped due to the end of the grant, and screening and diagnostic imaging was completed. A clinical follow-up of all randomized women is planned for at least 5 years.

## Preliminary results

From July 2013 to November 2015, a total of 1,254 women (mean age 47.2 ± 4.6 years) were enrolled: 624 were assigned to Mx plus US (standard of care arm) and 630 to MRI (experimental arm). The number of women recruited in each centre and their characteristics are reported in Table 2 and were balanced between the two study arms. Most women were below the age of 50 (72.2%) and premenopausal (44.6%) or perimenopausal (23.7%). More than one in four (27.6%) did not have children; 52.1% used oral contraceptives (currently or in the past); 15% of postmenopausal women had used HRT. Most women (86.3%) had a body mass index < 25. Only 4.6% of enrolled women (58/1254) had their first breast examination in this trial and most of them (44/58) were in their forties. Among women with a previous Mx, an extremely dense breast at the most recent Mx was recorded for 82.7% (984/1196). Breast and/or ovarian cancer in mother and/or sisters was reported by 36.5% of women (n=458) and 67 of them had a BC-LTR < 15%. A BC-LTR ranging from 15% to 30% was calculated for 41% (514/1254) and 47.5 % of them (244/514) had also a previous Mx classified as extremely dense. Overall, 59% of women (740/1254) had as sole inclusion criterion an extremely dense breast at Mx (LTR < 15% and BD = *d*).

**Table 2.** Recruitment centres, characteristics, and risk profiles of the 1,254 randomized women according to study arm

|  | **Mammography**<br>**plus Ultrasound** |  | **Magnetic**<br>**Resonance Imaging** |  |
| --- | --- | --- | --- | --- |
|  | **N.** | **%** | **N.** | **%** |
| **Total** | 624 | 100.0 | 630 | 100.0 |
| **Centres** |  |  |  |  |
| IRCCS Ospedale Policlinico San Martino, Genova | 139 | 22.3 | 139 | 22.1 |
| Ospedale Universitario Sant’Anna, Cona – Università degli Studi di Ferrara, Ferrara | 100 | 16.0 | 101 | 16.0 |
| Azienda Ospedaliera Universitaria Integrata, Verona | 100 | 16.0 | 100 | 15.9 |
| IRCCS Policlinico San Donato, San Donato Milanese | 53 | 8.5 | 54 | 8.6 |
| IRCCS Ospedale San Raffaele, Milano | 50 | 8.0 | 50 | 7.9 |
| Dipartimento di Scienze Radiologiche, Oncologiche, Patologiche – Università La Sapienza, Roma | 45 | 7.2 | 45 | 7.1 |
| Azienda Ospedaliera Universitaria “Santa Maria della Misericordia”, Udine | 39 | 6.2 | 39 | 6.2 |
| Fondazione IRCCS Istituto Nazionale dei Tumori, Milano | 34 | 5.4 | 38 | 6.0 |
| Fondazione Policlinico Universitario Agostino Gemelli IRCCS – Università Cattolica del Sacro Cuore, Roma | 34 | 5.4 | 34 | 5.4 |
| Istituto Nazionale Tumori IRCCS Fondazione G. Pascale, Napoli | 30 | 4.8 | 30 | 4.8 |
| **Age classes** |  |  |  |  |
| 40–44 | 214 | 34.3 | 206 | 32.7 |
| 45–49 | 236 | 37.8 | 249 | 39.5 |
| 50–54 | 130 | 20.8 | 121 | 19.2 |
| 55–59 | 44 | 7.1 | 54 | 8.6 |
| **Age at menarche (years)** |  |  |  |  |
| ≤ 11 | 118 | 18.9 | 141 | 22.4 |
| 12–13 | 336 | 53.8 | 311 | 49.4 |
| ≥ 14 | 165 | 26.4 | 171 | 27.1 |
| unknown | 5 | 0.8 | 7 | 1.1 |
| **Number of full-term pregnancies** |  |  |  |  |
| 0 | 170 | 27.2 | 176 | 27.9 |
| 1 | 208 | 45.8 | 176 | 38.8 |
| 2 | 211 | 46.5 | 231 | 50.9 |
| ≥ 3 | 35 | 7.7 | 47 | 10.4 |
| **Age at first birth (years)** |  |  |  |  |
| < 20 | 4 | 0.9 | 9 | 2.0 |
| 20–24 | 65 | 14.3 | 62 | 13.7 |
| 25–29 | 124 | 27.3 | 140 | 30.9 |
| 30–34 | 141 | 31.1 | 128 | 28.3 |
| ≥ 35 | 101 | 22.2 | 98 | 21.6 |
| Unknown | 19 | 4.2 | 16 | 3.5 |
| **Menopausal status** |  |  |  |  |
| Premenopausal | 282 | 45.2 | 277 | 44.0 |
| Perimenopausal | 150 | 24.0 | 147 | 23.3 |
| Postmenopausal | 117 | 18.8 | 130 | 20.6 |
| Unknown | 75 | 12.0 | 76 | 12.1 |
| **Contraceptive pill use** |  |  |  |  |
| Never | 303 | 48.6 | 298 | 47.3 |
| Current | 59 | 9.5 | 60 | 9.5 |
| Discontinued < 5 years | 39 | 6.2 | 46 | 7.3 |
| Discontinued $\geq 5$ years | 223 | 35.7 | 226 | 35.9 |
| **HRT use in postmenopause** |  |  |  |  |
| Never | 104 | 88.9 | 105 | 80.8 |
| Past | 9 | 7.7 | 21 | 16.1 |
| Current | 4 | 3.4 | 4 | 3.1 |
| **Body mass index (kg/m<sup>2</sup>)** |  |  |  |  |
| < 25 | 523 | 83.8 | 525 | 83.3 |
| 25–29 | 68 | 10.9 | 63 | 10.0 |
| $\geq 30$ | 12 | 1.9 | 17 | 2.7 |
| Unknown | 21 | 3.4 | 25 | 4.0 |
| **Cigarette smoking** |  |  |  |  |
| Never | 420 | 67.3 | 425 | 67.5 |
| Past | 107 | 17.1 | 97 | 15.4 |
| Current | 97 | 15.5 | 108 | 7.1 |
| **Alcohol consumption** |  |  |  |  |
| Never | 446 | 71.5 | 441 | 70.0 |
| Past | 39 | 6.2 | 41 | 6.5 |
| Current | 139 | 22.3 | 148 | 23.5 |
| **First degree family history of BC and OC** |  |  |  |  |
| None | 383 | 61.4% | 413 | 65.6% |
| BC | 218 | 34.9% | 197 | 31.3% |
| OC | 17 | 2.7% | 16 | 2.5% |
| BC and OC | 6 | 1.0% | 4 | 0.6% |
| **Breast density at pre-trial Mx** |  |  |  |  |
| A | 6 | 1.0 | 9 | 1.4 |
| B | 47 | 7.5 | 53 | 8.4 |
| C | 56 | 9.0 | 41 | 6.5 |
| D | 486 | 77.9 | 498 | 79.0 |
| Unknown | 29 | 4.6 | 29 | 4.6 |
| **LTR of BC** |  |  |  |  |
| < 15% | 358 | 57.4% | 382 | 60.6% |
| 15–30% | 266 | 42.6% | 248 | 39.4% |
| **Risk profile** |  |  |  |  |
| LTR ≥ 15%, breast density *a–c* | 109 | 17.5 | 103 | 16.3 |
| LTR < 15%, breast density *d* | 358 | 57.4 | 382 | 60.6 |
| LTR ≥ 15%, breast density *d* | 128 | 20.5 | 116 | 18.4 |
| LTR ≥ 15%, unknown breast density | 29 | 4.6 | 29 | 4.6 |

Mx, mammography; HRT, hormone replacement therapy; BC, breast cancer; OC, ovarian cancer; BD, breast density; LTR, lifetime risk.
Breast density at the pre-trial Mx: (*a*) almost entirely fat, (*b*) scattered fibroglandular densities, (*c*) heterogeneously dense, (*d*) extremely dense.

The distribution of major determinants of BC risk profiles of women enrolled in the study varied across centres: the rate of women recruited on the basis of an Mx classified as extremely dense ranged from 0% to 98.9% (Table 3).

**Table 3.**
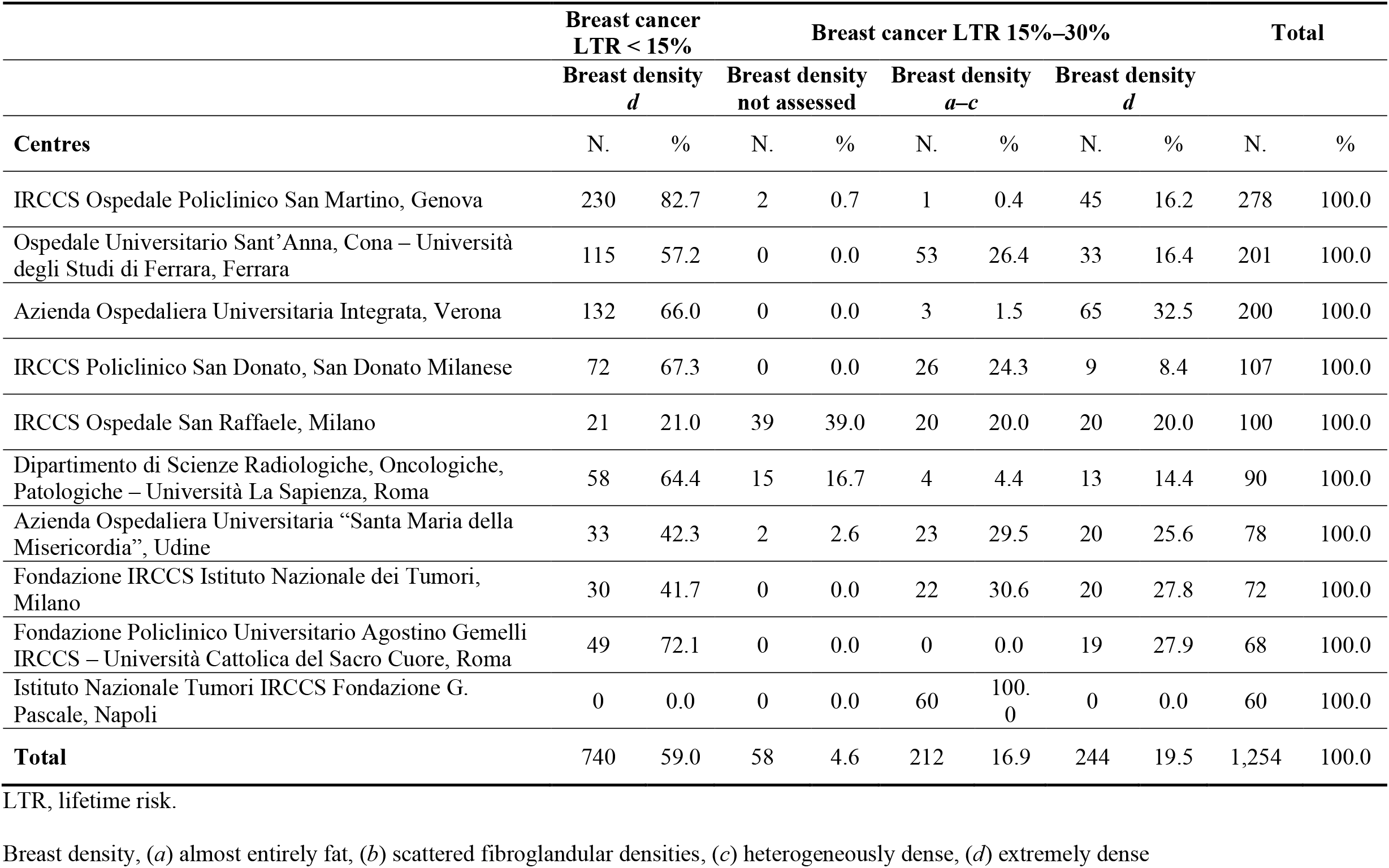
Distribution of the risk profiles of women enrolled in the 10 participating centres

|  | Breast cancer<br>LTR < 15% |  | Breast cancer LTR 15%–30% |  |  |  |  |  | Total |  |
| --- | --- | --- | --- | --- | --- | --- | --- | --- | --- | --- |
|  | Breast density<br>*d* |  | Breast density<br>not assessed |  | Breast density<br>*a–c* |  | Breast density<br>*d* |  |  |  |
| Centres | N. | % | N. | % | N. | % | N. | % | N. | % |
| IRCCS Ospedale Policlinico San Martino, Genova | 230 | 82.7 | 2 | 0.7 | 1 | 0.4 | 45 | 16.2 | 278 | 100.0 |
| Ospedale Universitario Sant’Anna, Cona – Università degli Studi di Ferrara, Ferrara | 115 | 57.2 | 0 | 0.0 | 53 | 26.4 | 33 | 16.4 | 201 | 100.0 |
| Azienda Ospedaliera Universitaria Integrata, Verona | 132 | 66.0 | 0 | 0.0 | 3 | 1.5 | 65 | 32.5 | 200 | 100.0 |
| IRCCS Policlinico San Donato, San Donato Milanese | 72 | 67.3 | 0 | 0.0 | 26 | 24.3 | 9 | 8.4 | 107 | 100.0 |
| IRCCS Ospedale San Raffaele, Milano | 21 | 21.0 | 39 | 39.0 | 20 | 20.0 | 20 | 20.0 | 100 | 100.0 |
| Dipartimento di Scienze Radiologiche, Oncologiche, Patologiche – Università La Sapienza, Roma | 58 | 64.4 | 15 | 16.7 | 4 | 4.4 | 13 | 14.4 | 90 | 100.0 |
| Azienda Ospedaliera Universitaria “Santa Maria della Misericordia”, Udine | 33 | 42.3 | 2 | 2.6 | 23 | 29.5 | 20 | 25.6 | 78 | 100.0 |
| Fondazione IRCCS Istituto Nazionale dei Tumori, Milano | 30 | 41.7 | 0 | 0.0 | 22 | 30.6 | 20 | 27.8 | 72 | 100.0 |
| Fondazione Policlinico Universitario Agostino Gemelli IRCCS – Università Cattolica del Sacro Cuore, Roma | 49 | 72.1 | 0 | 0.0 | 0 | 0.0 | 19 | 27.9 | 68 | 100.0 |
| Istituto Nazionale Tumori IRCCS Fondazione G. Pascale, Napoli | 0 | 0.0 | 0 | 0.0 | 60 | 100.0 | 0 | 0.0 | 60 | 100.0 |
| **Total** | 740 | 59.0 | 58 | 4.6 | 212 | 16.9 | 244 | 19.5 | 1,254 | 100.0 |

LTR, lifetime risk.
Breast density, (*a*) almost entirely fat, (*b*) scattered fibroglandular densities, (*c*) heterogeneously dense, (*d*) extremely dense

The frequency of women reporting one or more first-degree relatives affected by breast and/or ovarian cancer ranged from 26.6% to 91.7%.

## Discussion

In Italy, women aged 50 –69 years are offered biennial screening Mx independently of their BD and BC-LTR. Currently, an increasing number of women opt for regular surveillance imaging from age 40 onwards, particularly when they perceive being at increased B C risk [27]. Outside organized screening, women with high BD are usually offered yearly Mx plus US, even though data concerning the benefit of US supplemental screening in terms of reduced interval cancer rates are not consistent [20, 28] and even though supplemental US increases false positive findings [20, 29, 30]. In addition, it is not known whether the increased BC detection by US translates into reduced mortality.

Our study focuses on intermediate-risk women aged 40–59 years, only partially targeted by organized screening. To our knowledge, this is the first trial testing breast MRI as a stand-alone screening tool compared to Mx plus US. The rationale stemmed from observational high-risk studies showing a two-fold increase in MRI sensitivity compared to Mx/US, but without significant increase in sensitivity from the addition of Mx/US to MRI [14, 15, 31, 32]. The effect of an increased detection on BC-mortality and the entity of any MRI-associated overdiagnosis can be only addressed with RCTs, that are however difficult to conduct in high-risk settings due to ethical and psychological reasons. Therefore, two options are available: 1) to rely on ongoing and future uncontrolled studies, whose validity is undermined by biases affecting the comparison with external controls; 2) to conduct RCTs in women to whom MRI screening is not currently offered. The latter approach has two limitations: a) due to the lower BC risk, sample sizes will have to be much larger; b) the results will not be directly applicable to women to whom MRI screening is currently offered, due to different risk and possible different BC biology. However, these two options are not mutually exclusive, and both can give information on benefits, risks, harms, and costs associated with screening MRI.

Our population comprised healthy women with a 2 –3-fold increase in BC risk compared to a low-risk woman of the same age. BC-LTR was assessed with the IBIS Breast Cancer Risk Evaluation Tool version 6.0.0, that incorporates the most comprehensive set of personal risk factors and an extensive family history of breast and ovarian cancer. Since the IBIS Breast Cancer Risk Evaluation Tool does not integrate BD, we extreme BD as an eligibility criterion. Over 70% of women recruited were in their forties, the age range in which screening recommendations are not consistent across Europe [33]. Due to recruitment at radiology units, 95% of enrolled women had a pre-trial Mx, but the distribution of risk profiles varied widely across centres (from 0% to 99% of women having extremely dense breasts), showing different ultimate sources of recruitment (e.g., self-referral for breast examination or collaboration with familial cancer clinics for the surveillance of women at increased BC risk who did not carry pathogenic germ-line variants). In our study, around one in four women with extreme BD had also a BC-LTR ≥ 15%: in this subgroup a BC-LTR recalculation using the 2017 IBIS Breast Cancer Risk Evaluation Tool version 8b that includes BD in the model might take the risk over the 30% threshold in a high number of cases.

Our study has some limitations: it was designed as a preliminary, feasibility study with the goal of recruiting 2,000 women in 2.5 years, but in the designed period we recruited only 1,254 women. Each centre was required to enrol 200 women, but only three out of ten centres reached the target. We observed that the study budget did not adequately incorporate staff needed to support such a study; for instance, significant workflow demands fell on the radiologists, as one of the required assessments for eligibility was the evaluation of previous Mx with subsequent BC-LTR calculation. Time constraints and concurrent competing trials were causes of the failure to reach the expected recruitment. As in other trials, inadequate funding and complexity of the study design were the reasons that contributed to unsuccessful trial recruitment [34]. Moreover, when dealing with first-round MRI there we can face a high rate of BI-RADS 3 designations. Especially in the case of only intermediate risk population (greatly lowering the pre-test cancer probability in comparison with *BRCA* or *TP53* mutation carriers) and in the absence of any correlate at reassessed Mx and targeted US, we hypothesized that the residual cancer rate could be sufficiently low to postpone the MRI to the year after. This approach has been already investigated by Elshof et al. [35] for additional MRI-detected lesions outside the primary tumour region in the preoperative setting, where the pre-test cancer probability should be higher than in any screening setting. Additional lesions outside the primary tumour region without any correlate at targeted US were found in 81 out of 690 patients. None of them resulted in malignant disease at follow-up after breast conserving therapy (mean follow-up time: 57.1 months). To minimize the risk of a diagnostic delay, we also planned a 3-month follow-up with Mx and US. Furthermore, we highlight that our intermediate screening setting implied an expected low cancer detection rate: researchers had to reckon with this expectation, trying to minimize unnecessary biopsies and overdiagnosis [13, 36– 38].

Despite these limitations, the present trial will give useful information on acceptability of the two screening models, on the risk profile of women who accepted to participate, on the diagnostic performance in terms of second-look examinations, short-term re-evaluation, invasive procedures, and diagnostic yield. The planned 5-year follow-up should provide an estimate of the magnitude of overdiagnosis, if any, associated to MRI.

Patient compliance remains a major issue in breast MRI screening [39]: Berg et al. [40] reported that over 40% of women at high BC risk refused to undergo additional MRI screening; a similar result was observed in the DENSE trial [24]. In addition, MRI screening is associated with high costs: instrumental, technical (contrast agent, acquisition time, post-processing), and regarding interpretation time. In the last years, new approaches were investigated to decrease breast MRI costs and to make breast MRI more accessible. Abbreviated protocols reducing acquisition and interpretation times demonstrated an accuracy equivalent to that of full protocols [41–43]. This approach could reduce the costs of breast MRI, making it accessible to a broader population and thus potentially increase women compliance.

## Data Availability

This article details the study design and characteristics of patients enrolled in the "MRIB" multicentre randomized controlled trial. All remaining study data will be the object of a separate publication.

## Abbreviations

ACR: American College of Radiology;
BC: breast cancer
BD: breast density;
HRT: postmenopausal hormone replacement therapy;
MRI: contrast-enhanced magnetic resonance imaging;
Mx: mammography;
RCT: randomized controlled trial
US: breast ultrasound.

